# Acoustic Analysis of Primary Care Patient-provider Conversations to Screen for Cognitive Impairment

**DOI:** 10.64898/2025.12.27.25343088

**Authors:** Joseph T. Colonel, Jacqueline Becker, Lili Chan, Cara Faherty, Tielman van Vleck, Laura Curtis, Juan Wisnivesky, Alex Federman, Baihan Lin

**Affiliations:** Department of Psychiatry, Icahn School of Medicine at Mount Sinai, New York, NY, USA; Division of General Internal Medicine, Icahn School of Medicine at Mount Sinai, New York, NY, USA; Institute for Personalized Medicine, Icahn School of Medicine at Mount Sinai, New York, NY, USA; Division of General Internal Medicine, Feinberg School of Medicine at Northwestern University, Chicago, IL, USA

**Author notes:** Corresponding Author: Joseph T Colonel, PhD Icahn School of Medicine at Mount Sinai 3 East 101st Street, New York, NY 10029. co-senior authors (Alex Federman, Baihan Lin).

## Abstract

**Importance:** Cognitive impairment (CI) is often under detected in primary care due to time and resource constraints. Passive analysis of clinical dialogue may offer an accessible approach for screening.

**Objective:** To assess whether audio recordings of patient–physician dialogue during routine primary care visits can be used to identify CI using acoustic speech features and machine learning (ML).

**Design:** This observational study conducted among older primary care patients involved audio recording primary care visits using a microphone and portable device. An external validation cohort was recruited in a separate city to assess reproducibility of findings.

**Setting:** The study was conducted in primary care practices in New York City, with additional participants recruited from primary care practices in Chicago, Illinois, for validation.

**Participants:** The study included 787 English-speaking patients aged 55 years and older, without documented history of dementia or mild CI. Eligible patients were recruited from primary care practices during routine visits. For validation, 179 patients meeting the same eligibility criteria were recruited from primary care practices in Chicago.

**Exposures:** Multiple thirty-second speech segments were extracted from recordings. Acoustic features were derived using foundation models (Whisper, HuBERT, Wav2Vec 2.0) and expert-defined methods (eGeMAPS, prosody).

**Main Outcomes and Measures:** CI was defined as Montreal Cognitive Assessment score ≥1.0 standard deviations below age and education-adjusted norms. ML classifiers were trained to predict CI status from audio recordings. We calculated area under the receiver operating characteristic curve (AUC-ROC) and maximum F1 score (F_max_) for identifying CI participants.

**Results:** The mean age was 66.8 years and 21% had CI. Models using Whisper-derived acoustic features performed best (AUC-ROC=0.733, 95% confidence interval [95%CI]=0.714-0.752; F_max(CI)_=0.504, 95%CI=0.474-0.534). Results generalized to the external site with similar performance (AUC-ROC=0.727, 95%CI=0.714-0.740; F_max(CI)_=0.459, 95%CI=0.442-0.476).

Model interpretation identified pitch, timing, and variability features as key predictors. When used for screening, the algorithm achieved positive predictive value of 30.4% (95%CI=28.7%-32.1%), sensitivity of 68.2% (95%CI=61.8%-74.6%), and specificity of 63.6% (95%CI=59.8%-67.4%) on the holdout cohort.

**Conclusions and Relevance:** ML models trained on acoustic features from brief clinical conversations identified CI with high accuracy. These findings support the feasibility of passive, speech-based screening during routine primary care.

**Key Points:** 

**Question:** Can acoustic features extracted from audio recordings of patient–physician conversations during routine primary care visits be used to screen for cognitive impairment?

**Findings:** In this study including 787 older adults without diagnosis of cognitive problems, machine learning models trained on acoustic features from audio segments of recordings of primary care visits achieved area under the receiver operating characteristic curve values of 0.72 for predicting cognitive impairment. The algorithm achieved a sensitivity of 83%, specificity of 44%, and positive predictive value of 28%, identifying a subset of primary care patients at higher risk for cognitive impairment. Models performed similarly on an external validation dataset of 179 participants. Interpretability analyses highlighted patient pause duration and energy-related features as salient indicators of cognition status.

**Meaning:** These findings suggest that short segments of naturalistic clinical dialogue may contain useful acoustic signals for passively screening patients for cognitive impairment.

## Introduction

Cognitive impairment (CI) affects nearly 1 in 5 adults in the United States aged 65 years and older,^1^ yet it often goes unrecognized by clinicians until it reaches advanced stages. ^1–8^ It is also less likely to be recognized in minoritized older adults, potentially furthering disparities in health care delivery and outcomes.^9^ The United States Preventive Services Task Force has found insufficient evidence to assess the balance of benefits and harms for early detection of CI through screening.^10^ Nevertheless, there are various reasons for screening and early detection, including identifying and addressing reversible causes of CI, reducing risks of harm that may be caused by medications, addressing co-morbid conditions (e.g., depression) that may worsen cognitive functioning, improving support for caregivers to alleviate the stress and burden associated with caring for people with CI,^11–19^ and improving self-management support^20^ and coordination of care.^18,21^ Additionally, new treatments for Alzheimer’s Disease (AD) have recently emerged that can slow cognitive decline in dementia and the benefits may be greatest for those treated earlier in the disease course. ^22–23^

Primary care is the most appropriate setting for dementia screening.^1,4,7,24^ However, routine dementia screening occurs infrequently in primary care despite the availability of brief assessments like the Mini-Mental State Examination (MMSE) and the Montreal Cognitive Assessment (MoCA).^2,5,25^ Overall, fewer than one-third of older adults are ever screened for CI.^1,2,26^ Instead, primary care providers typically assess cognitive functioning when patients present with a specific cognitive complaint. Moreover, they miss CI in up to 76% of patients when using routine data gathering (e.g., history, physical examination).^5^ Competing priorities for primary care physicians, like providing routine preventive care, may contribute to the low rates of CI screening in primary care settings.

To overcome some of the barriers to screening, researchers have worked to develop automated CI detection using machine and deep learning models applied to medical record data.^27–31^ Such models have performed well for identifying patients with known CI,^32^ but are not accurately performing in screening for new cases. ^28^ A promising new avenue for automated CI detection is the analysis of speech, which is increasingly recognized as a clinical biomarker of cognitive functioning.^33–34^ Low-cost, high-quality voice recorders saturate the market, ^35^ greatly expanding the feasibility of speech-based CI screening in clinical settings.

Beyond laboratory-based assessments and electronic health record approaches, there has been growing interest in moving CI prediction closer to the point of care. Prior studies have explored the use of short, structured speech tasks (e.g., picture description) administered in research settings to train machine learning models for Alzheimer’s disease and MCI detection. ^36^ While these approaches demonstrate strong diagnostic accuracy, they often require specialized test administration, standardized recording environments, and substantial participant time, which limit scalability and generalizability. In contrast, fewer studies have examined speech generated during naturalistic clinical encounters.^37^

While these and other studies demonstrate proof of concept, their application to CI screening in primary care is limited by design. Specifically, none were designed to identify individuals in primary care with no prior history of CI. In addition, embedding speech-based CI screening into existing care processes could overcome key barriers of cost, time, and accessibility, while capturing ecologically valid representations of patients’ everyday communicative and cognitive functioning. Our study extends beyond controlled laboratory contexts by evaluating speech from real-world primary care encounters. We recruited patients without known CI, administered a validated cognitive screener as a gold standard, and developed machine learning algorithms for CI screening in primary care settings.

## Methods

### Subjects and Settings

Patients were recruited from primary care practices in the Mount Sinai Health System in New York City, NY and the Northwestern Memorial Hospital in Chicago, IL from August 2020 through December 2021. The practices included three hospital-based teaching clinics and two multi-provider faculty clinics. We included patients attending a routine primary care visit who were ages 55 and older and spoke English. Patients with a diagnosis of MCI, dementia or related disorders in the problem list or medical history of their medical record were excluded (ICD10-CN codes G20, G30, G31, G91, F00-F03, F09, R41, R47, I69). Study procedures were approved by the Institutional Review Boards of the Icahn School of Medicine at Mount Sinai and the Northwestern University Feinberg School of Medicine.

### Recruitment and Interviews

Research coordinators (RC) screened a list of potentially eligible patients scheduled for a routine primary care visit generated from the health systems’ electronic medical records and obtained permission of the patients’ primary care providers to recruit them for study participation. A recruitment letter was then mailed to a random selection of individuals followed by a telephone call, during which the RC described the study and administered an eligibility screener. Subjects meeting final eligibility criteria were invited for an in-person interview scheduled to immediately follow their upcoming primary care appointment. The 30-minute interviews were conducted by trained RCs in an exam room.

### Audio Data Collection

Participants were recorded during their scheduled visits with primary care physicians using a Tascam DR-10L Micro Portable Audio Recorder with lavalier attached to a lapel or collar at a 44.1 kHz sampling rate, stored as 16-bit linear pulse code modulation (PCM) waveform audio (WAV) files.

### Gold Standard Assessment of Cognitive Impairment

We used the MoCA as the gold standard assessment of CI because it is widely used for dementia screening.^9, 38–40^ It consists of 12 tasks covering visuospatial/executive functioning, naming, memory, attention, language, delayed recall and orientation. Scores range from 0-30. Raw scores were converted to age and education adjusted z-scores. We then defined CI as MoCA scores ≥1.0 standard deviation below the mean of normative data. ^39,41–44^ A detailed analysis of CI in the study cohort is described elsewhere.^3^

Data were also collected on standard sociodemographic characteristics of study participants, including age, gender, race and ethnicity, English language proficiency (ELP) and country of birth. ELP was assessed with a single item, “How would you describe your ability to speak and understand English?” with 6 response options ranging from very poor to excellent.

Low ELP was defined as a response of very poor, poor or fair.

## Acoustic Methods

### Acoustic Preprocessing

We evaluated several preprocessing pipelines to assess how acoustic processing influences classification performance and model generalizability. Analyses were conducted on two audio types: recordings containing both patient and clinician speech, and recordings containing only patient speech.

For the combined audio, we tested untreated recordings and versions processed with voice activity detection (VAD) to reduce noise using Silero VAD.^45^ VAD was applied to the full-length recordings to identify dialog passages. To control potentially confounding acoustic noise, all audio segments with no detected voice were silenced. All others were unaltered to preserve the dialog’s acoustic characteristics.

For patient-only audio, we applied the Cocktail Fork Separation (CFS) algorithm to isolate the patient’s voice and silence background noise and clinician speech. ^46^

### Acoustic Feature Extraction

We extracted five sets of acoustic features that capture foundation model, paralinguistic, and prosodic information from patient speech. Deep neural network (DNN) feature extraction was performed on mono audio downsampled to 16 kHz for compatibility with pre-trained models (Whisper, ^47–48^ HuBERT, ^49–50^ Wav2Vec2.0^51–52^) by extracting and averaging final-layer embeddings. Expert-defined feature extraction (eGeMAPS, ^53–55^ prosody^56–58^) was performed on the 44.1kHz mono waveform. Features were extracted on non-overlapping 30 second segments of each recording. Full details regarding each featureset can be found in the Supplementary materials.

## Machine Learning Methods

### Machine Learning Setup

To predict CI from spontaneous patient speech, we implemented a two-stage machine learning framework: (1) segment-level classification of speech excerpts, and (2) participant-level

aggregation of segment predictions. The design was optimized for robustness to class imbalance and interpretability in clinical contexts. A diagram of our full pipeline is shown in Figure 1.

**Figure 1.**
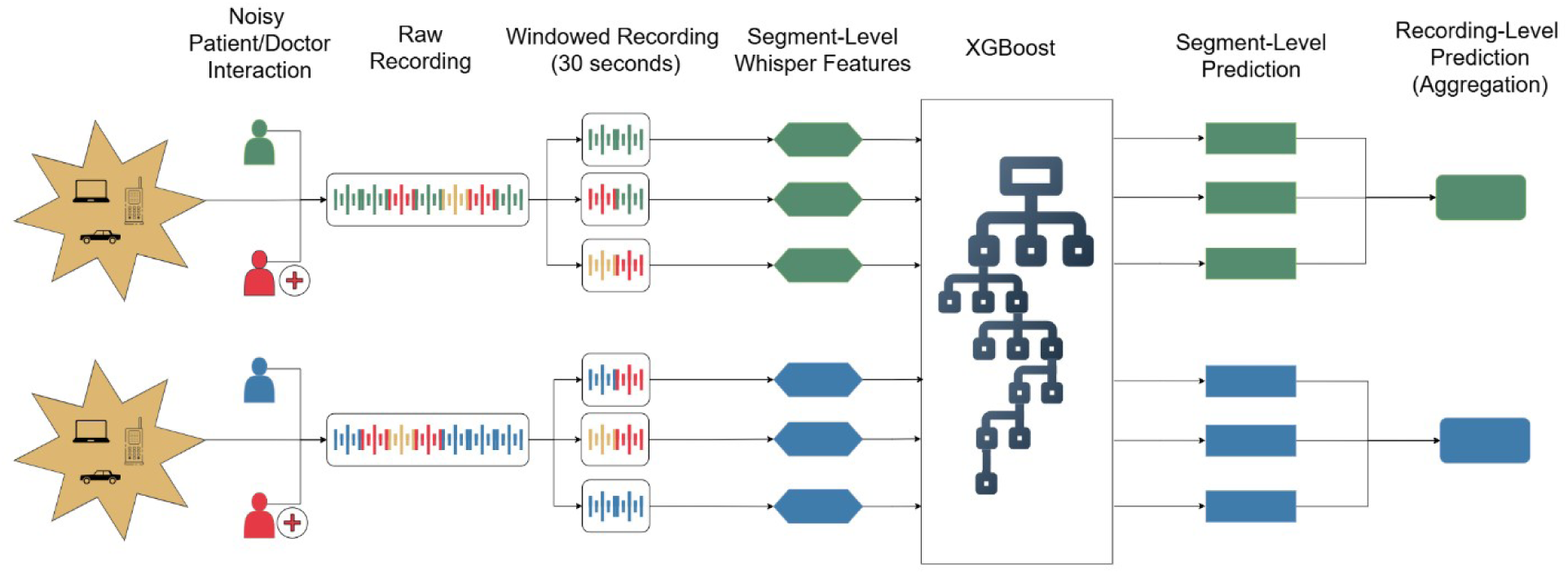
Acoustic preprocessing, feature extraction, and classification pipeline.

### Segment-Level Processing

Each participant’s speech was divided into fixed, non-overlapping 30-second segments. All acoustic features (Whisper, HuBERT, W2V2, eGeMAPS, and prosody) were extracted per segment. Feature vectors were z-normalized using statistics computed on the training folds during cross-validation.

### Segment-Level Classification

We trained an XGBoost classifier to predict whether each segment was associated with CI.^59^ One classifier was trained per feature set. Due to class imbalance at the participant level, we applied random oversampling of the minority class (cognitively impaired participants) in the training set. Oversampling was performed at the participant level, duplicating all segments from selected participants to preserve intra-subject structure and avoid data leakage.

### Participant-Level Aggregation

Participant-level predictions were generated by aggregating segment-level impairment probabilities. We evaluated several aggregation strategies including mean, median, and BioBERT inspired aggregation,^60^ where we upweighted the segment with the highest predicted probability by a factor of c relative to the mean of the remaining segments. We evaluated values of c ∈ {1, 2, …, 9}, optimizing c via cross-validation.

### Model Evaluation

Model performance was evaluated using k-fold cross-validation stratified by CI prevalence at the participant level to ensure no leakage of speech content across folds. We report Area Under the ROC Curve (AUC-ROC) and F_max_ of the CI class (F_max(CI)_),^61^ which is defined as maximum F1 score achieved across all probability thresholds, computed specifically for the CI class. All metrics were averaged across folds.

To evaluate generalizability, we assessed model performance on an external holdout dataset collected at Northwestern Memorial Hospital clinics using the same training folds derived from the Mount Sinai data.

### Positive Predictive Values for Clinical Application

To assess potential clinical applicability, we explored CI classifiers built using the Whisper-derived speech embeddings from the Mount Sinai cohort. Each model configuration corresponded to a different preprocessing pipeline and was evaluated using AUC-ROC to identify the best-performing classifier. This selected classifier formed the basis of a speech-based screening algorithm designed to flag individuals for formal cognitive assessment based on their recorded clinical dialogue.

For screening implementation, each patient’s dialogue recording was processed through the trained classifier, which produced a continuous output between 0 and 1 representing the predicted probability of CI at the end of the recording. A predefined threshold was then applied to this probability such that when the estimated probability of CI was higher than the threshold, the patient would be flagged for further CI screening. Three preprocessing pipelines were used, each requiring a distinct threshold (0.25 for the raw, 0.39 for the VAD, and 0.28 for the CFS audio).

Thresholds were chosen to prioritize positive predictive value (PPV), sensitivity, and specificity, as these measures are more clinically relevant than overall classification accuracy in screening contexts. Assuming a 20% prevalence of undiagnosed CI in primary care, ^3^ consistent with prior estimates, we selected thresholds to achieve a target PPV of approximately 30%, balancing clinical utility with acceptable false-positive rates.

## Results

### Study Enrollment and Participant Characteristics

Among 3,815 potentially eligible persons, RCs contacted 2,894 (75.9%) by telephone; 1,737 (60.0%) declined study participation and 208 (7.2%) were unable to complete screening in English; 949 (32.8%) were eligible and agreed to study participation, 918 provided signed consent and 787 (20.1%) completed the in-person interview and had a primary care dialogue recorded.

The mean age of study participants was 66.8 years; 55% were male, 32.4% Black, and 25.7% were Hispanic (Table 1). The overall rate of undiagnosed CI based the MoCA was 20.7%.

### Collected Audio Data

At the Mount Sinai clinics, a total of 388.6 hours of audio were recorded, with mean [SD] primary care encounter duration of 29.6 [13.0] minutes. At the Northwestern clinics, a total of 70.6 hours of audio were recorded, with mean primary care encounter duration of 23.7 [9.7] minutes. Demographic characteristics of the study populations stratified by site are shown in Supplemental Table 1.

### Classification Performance by Acoustic Feature Set

Performance of the different models for classification in the derivation set are shown in Figure 2. The DNN-derived features outperformed the expert-defined features in the evaluation. Whisper embeddings achieved the highest overall classification performance, achieving a mean AUC-ROC of 0.733 (95% confidence interval [95% CI] = 0.714-0.752) and mean F_max(CI)_ of 0.502 (95% CI = 0.471-0.533) on the unmodified audio recordings, followed by HuBERT-derived and W2V2 within the DNN-based features (See Figure 2).

**Figure 2.**
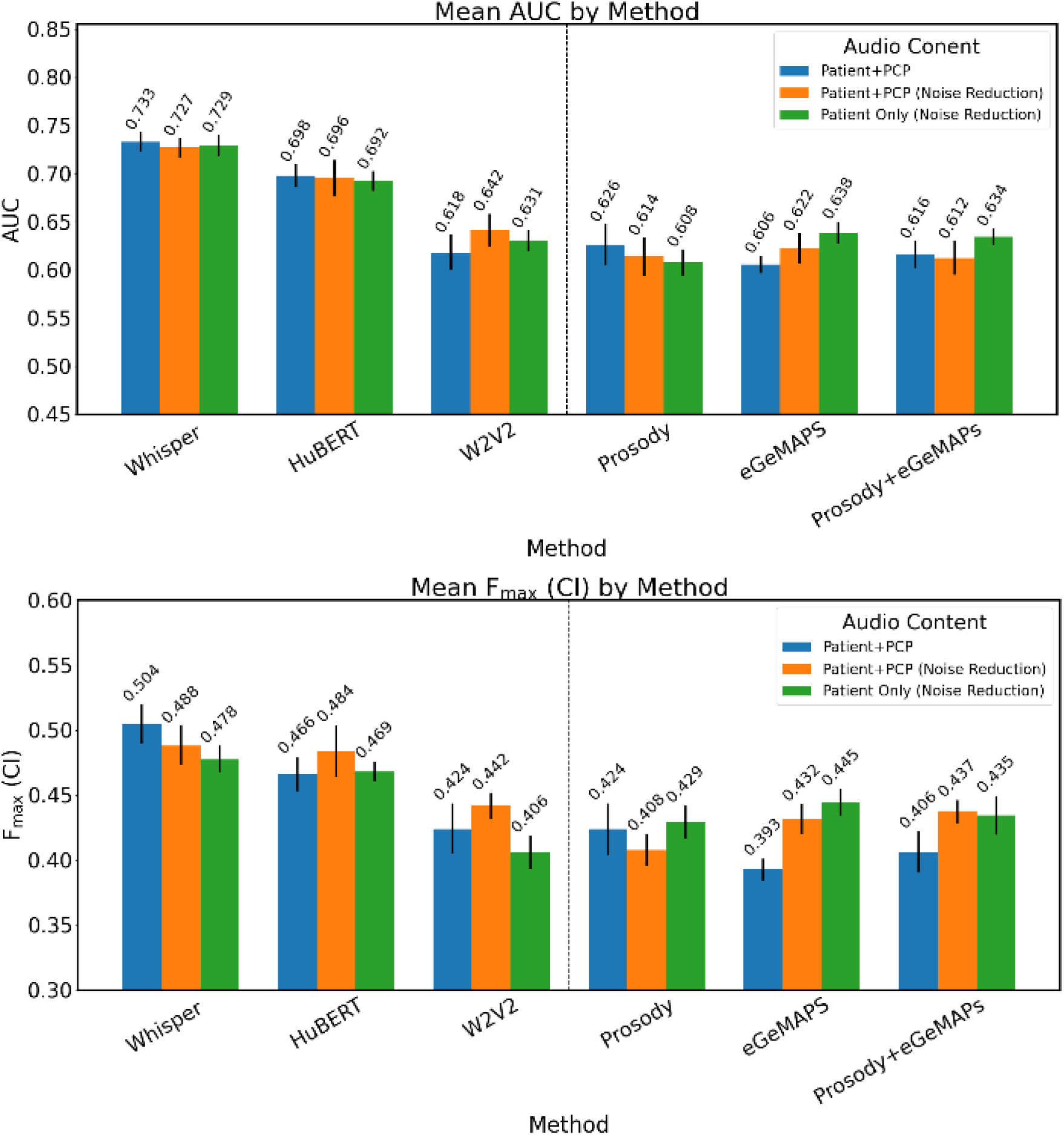
Classification performance on derivation dataset (N=787) n.

Models based on eGeMAPS and prosodic features showed lower but non-negligible performance. Of the two featuresets, eGeMAPS performed best on isolated patient audio, with mean AUC-ROC of 0.638 (95% CI = 0.615-0.661) and F_max(CI)_ of 0.445 (95% CI = 0.426-0.464), with the prosody features performing slightly better on the unmodified audio.

### External Validation

Results of the validation models are shown in Figure 3. Models trained on Whisper-derived features demonstrated the highest generalizability, maintaining comparable classification performance relative to internal testing. The best performing model achieved mean AUC-ROC of 0.727 (95% CI = 0.714-0.740) on noise-reduced dialogue audio and F_max(CI)_ of 0.459 (95% CI = 0.441-0.477) on untreated audio. Similarly, models utilizing prosodic features from patient-only audio segments showed stable performance, with mean AUC-ROC of 0.607 (95% CI = 0.596-0.618) and F_max(CI)_ of 0.363 (95% CI = 0.349-0.377). In contrast, all other models had diminished predictive accuracy when applied to the external dataset.

**Figure 3.**
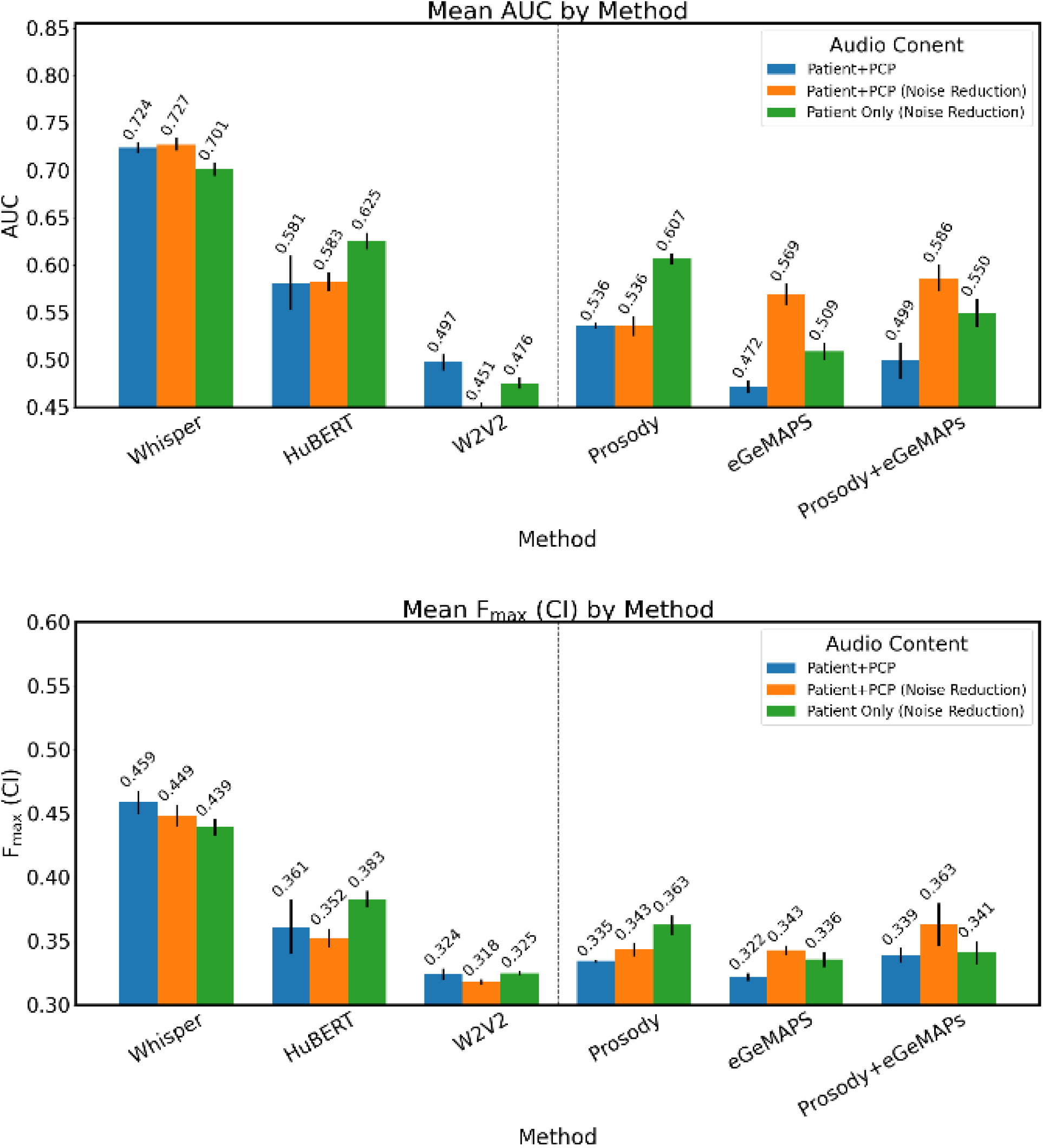
Classification Performance on evaluation dataset (N=179)

### Screening Algorithm

The Whisper model employing data with noise reduction preprocessing had the best performance for screening, with sensitivity of 83.4% (95% CI = 79.2%-87.6%) and specificity of 44.4% (95% CI = 42.1%-46.7%) in the derivation set (at PPV 28.2% (95% CI = 26.4%-29.6%)) and sensitivity of 68.2% (95% CI = 61.8%-74.6%) and specificity of 63.6% (95% CI = 59.8%- 67.4%) in the validation set (at PPV 30.4% (95% CI = 28.7%-32.1%)) (Table 2). Sensitivity was lower in the validation set with raw audio and CFS preprocessing.

**Table 2.** Performance of Screener on Derivation and Validation datasets ± 95% Confidence.

| Preprocessing | AUC-ROC | Positive Predictive Value | Sensitivity | Specificity |
| --- | --- | --- | --- | --- |
| Raw Audio |  |  |  |  |
| Derivation set | 0.733±0.020 | 31.1±2.3% | 82.1±4.5% | 52.4±3.1% |
| Validation set | 0.724±0.011 | 30.4±0.9% | 51.7±7.4% | 72.6±3.0% |
| Noise Reduction |  |  |  |  |
| Derivation set | 0.727±0.019 | 28.2±1.8% | 83.4±4.2% | 44.4±2.3% |
| Validation set | 0.727±0.013 | 30.4±1.7% | 68.2±6.4% | 63.6±3.8% |
| Source Separation |  |  |  |  |
| Derivation set | 0.729±0.021 | 30.7±1.6% | 83.9±4.2% | 50.6±2.0% |
| Validation set | 0.701±0.014 | 30.4±1.8% | 49.4±4.2% | 73.6±2.4% |

### Model Interpretation

Model interpretability analyses provided insights into the specific speech characteristics associated with CI. Using SHAP (Shapley Additive Explanations)^62^ values applied to the XGBoost models trained on prosodic features derived from patient-only audio, the top 10 influential features for prediction are plotted in Figure 4. Increased energy in unvoiced segments and greater variability in pause duration were associated with higher likelihood of predicting CI. In contrast, higher values of features related to fundamental frequency (F0), voicing rate, and energy dynamics were more indicative of predicting healthy cognition.

**Figure 4.**
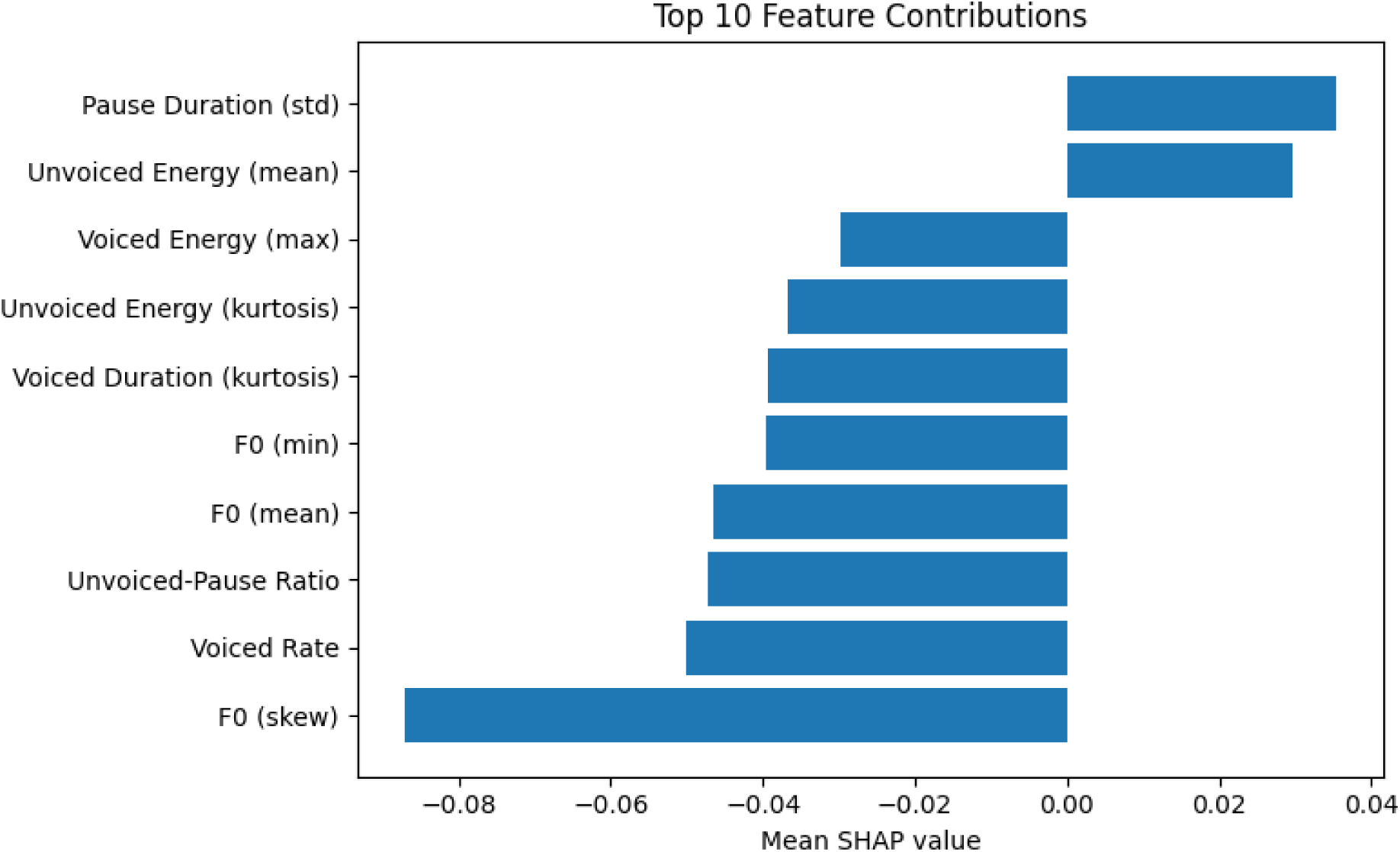
SHAP interpretation analysis of prosody featureset on Northwestern holdout data.

## Discussion

We developed and evaluated machine learning models using acoustic foundation model embeddings derived from primary care dialogue. Among several architectures tested, models based on Whisper embeddings achieved the highest predictive accuracy, with mean AUC-ROC values exceeding 0.72 in a holdout dataset. After setting a screening cutoff threshold for the output of the classification model, the algorithm yielded sensitivity of 68.2% and specificity of 63.6% in the validation set (at PPV 30.4%). Preprocessing techniques that reduced ambient noise while preserving conversational integrity yielded the most robust performance on holdout data.

These findings highlight both the feasibility and technical promise of a scalable, speech-based cognitive assessment for primary care. By leveraging naturally occurring speech from primary care visits, and without adding burden to clinical workflows, our approach could complement existing screening strategies and support earlier detection of CI in real-world care settings.

To our knowledge, this is the first study to conduct passive screening for undiagnosed CI from recorded patient-clinician dialog, and the classification performance aligns with screening literature using other stimuli. For example, Schäfer et al ^31^ achieved AUC-ROC of 0.73-0.77 on intra-cohort CI screening when evaluating a speech-based biomarker for CI. In contrast to our assessment of cognition under naturalistic conditions, the Schäfer study used a mobile recording platform that recorded participants undergoing formal cognitive assessments like semantic verbal fluency. Our findings highlight the potential of unstructured conversational data as a novel and complementary modality for passive CI screening.

The generalizability of the Whisper-based models, as evidenced by their consistent performance on the holdout data, may be due to the diverse acoustic conditions and speaker demographics seen in the web-scale, multilingual dataset used to train the foundation model. In comparison, HuBERT and W2V2 were primarily trained on datasets of English-language audiobook recordings. Similarly, prosodic features may have outperformed the eGeMAPS features as prosodic measures are more robust to varying recording environments than eGeMAPS features.^63^

These findings suggest that features from speech foundation models may capture clinically relevant patterns more effectively than traditional acoustic features. This observation aligns with state-of-the-art acoustic approaches for CI classification from structured laboratory assessments, where winners of the PROCESS^36^ and TAUKADIAL challenges^64^ used Whisper-derived acoustic features for classification.^48,65^

The tradeoff of using DNN-based features is reduced interpretability. Alternatively, expert-defined features, though underperforming, provide interpretable insights into physiological changes associated with CI since they explicitly derive from speech production processes. Therefore, we performed interpretation analysis on the best expert-defined feature model. These findings suggest that disrupted speech timing, reduced vocal dynamism, and sex-related pitch characteristics contribute to the acoustic profile of CI. This agrees with previous findings showing pitch-related prosody features vary significantly with CI in Parkinson’s disease.^66^ Other meta-analyses have highlighted that pause-related features and rate of speech vary significantly between CI and control populations.^67–68^

### Limitations

This study has several limitations. Although recordings were collected during routine care, all data were collected in two affiliated clinical sites using a uniform protocol, which may limit generalizability. Second, while the models capture differences in acoustic features, they do not account for lexical content, which may provide complementary diagnostic value. Third, the inclusion of only English speakers may limit the applicability of this approach to non-English speaking populations.

## Conclusion

This study shows that analyzing the acoustic patterns of ordinary patient–clinician conversations could help fill an important gap in cognitive screening. Machine learning models trained on acoustic features from brief clinical conversations identified CI with high performance in-line with the CI screening literature. These findings support the feasibility of passive, speech-based screening during routine primary care. Future work should validate these findings in larger and more diverse populations, assess longitudinal predictive utility, and explore integration with existing electronic health record data. Ultimately, speech-based screening tools may offer a noninvasive, low-cost approach to identifying CI earlier and more equitably across clinical populations.

## Data Availability

Data available:
Yes
Data types:
Other (please specify)
The datasets generated and analyzed during the current study are available from the corresponding author on reasonable request.
How to access data:
When available:
With publication
Document types:
None
Who can access the data:
The datasets generated and analyzed during the current study are available from the corresponding author on reasonable request.
Types of analyses:
Data will be made available for specific requests on a case-by-case basis.
Mechanisms of data availability:
With a signed data sharing agreement.

## Acknowledgments

This work was supported in part by Award Number R01AG066471 from the National Institute Aging of the National Institutes of Health.

This work was also supported in part through the Minerva computational and data resources and staff expertise provided by Scientific Computing and Data at the Icahn School of Medicine at Mount Sinai and supported by the Clinical and Translational Science Awards (CTSA) grant UL1TR004419 from the National Center for Advancing Translational Sciences. Research reported in this publication was also supported by the Office of Research Infrastructure of the National Institutes of Health under award number S10OD026880 and S10OD030463. The content is solely the responsibility of the authors and does not necessarily represent the official views of the National Institutes of Health.

Dr. Wisnivesky received consulting honorarium from Sanofi, Banook, PPD, and AMA and research grants from Sanofi, Regeneron, Axella, and Boehringer Ingelheim.

## SUPPLEMENTARY MATERIALS

**Supplemental Table 1.**
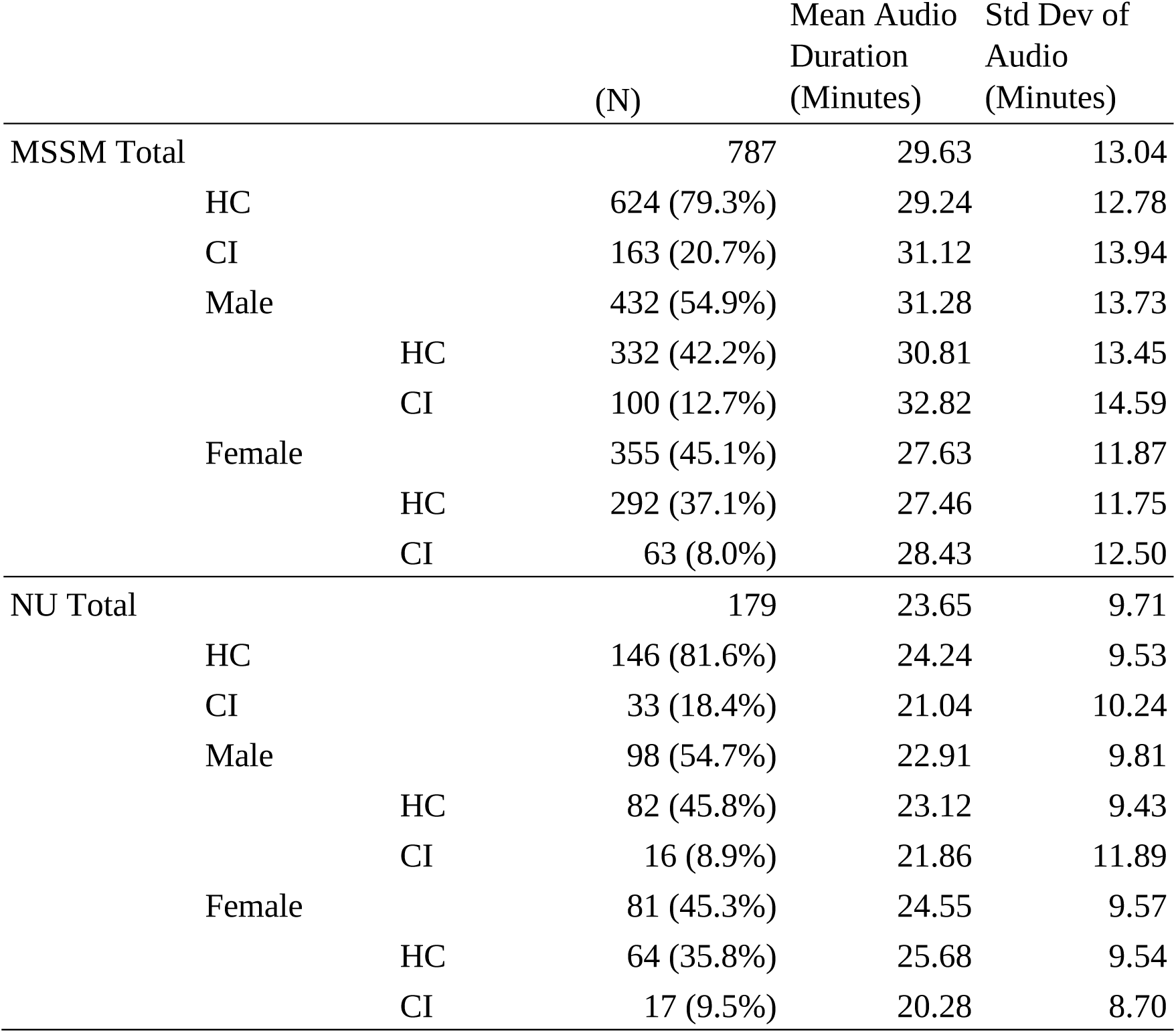
Mount Sinai and Northwestern Audio Data.

## Supplemental Section 1. Acoustic Feature Extraction

*Whisper Embeddings*: We used the audio encoder from OpenAI’s Whisper large-v2 model to derive high-level acoustic embeddings from each 30-second segment. ^47^ The model’s multilingual training and robustness to real-world noise make it well-suited for clinical settings. ^48^

*HuBERT Embeddings*: We applied the Hidden-Unit BERT (HuBERT) Base model (finetuned on LibriSpeech) to generate self-supervised acoustic representations. ^49^ HuBERT clusters latent speech units and uses them to predict masked inputs, capturing subphonemic patterns relevant for cognitive and affective speech analysis.^50^

*Wav2Vec 2.0 Embeddings*: We extracted representations from the Wav2Vec 2.0 Base Robust model (W2V2).^51^ Like HuBERT, W2V2 learns contextual embeddings using contrastive predictive coding. These embeddings offer complementary information regarding speech fluency, articulation, and rhythm.^52^

*eGeMAPS Acoustic Features*: We computed the Extended Geneva Minimalistic Acoustic Parameter Set (eGeMAPS) using the openSMILE toolkit.^53–54^ eGeMAPS includes 88 low-level descriptors such as jitter, shimmer, formant frequencies, MFCCs, and spectral slope. These features have been linked to neurological and psychiatric status. ^55^

*Prosodic Features*: Prosodic cues including pitch (F0), speaking rate, pause duration, and voicing probability were extracted using the DisVoice toolkit.^56–57^ These features quantify the suprasegmental aspects of speech and have been shown to vary in cognitive decline. ^58^

